# Longitudinal and comparative analysis of humoral response upon COVID-19 vaccination

**DOI:** 10.1101/2021.10.14.21264762

**Authors:** Salvador Romero-Pinedo, Marina Quesada, Stela Álvarez-Fernández, Asunción Olmo, David Abia, Balbino Alarcón, Pilar Delgado

## Abstract

The emergence of COVID-19 has led to a worldwide challenge for the rapid development of vaccines. Several types of safe and effective vaccines have been available in a time frame never seen before. Comparative studies to know the extent of protection and the immune response elicited by the different vaccines are of outstanding utility. Here, as a correlate for protection, we perform a comparative study of the humoral response to three vaccines, ChAdOx1 (Oxford-AstraZeneca), mRNA-1273 (Moderna), and BNT162b2 (Pfizer-BioNTech) by applying a flow cytometry-based highly sensitive method that we had previously developed. We have found that mRNA vaccines (mRNA-1273 and BNT162b2) induce a stronger humoral response that lasts for at least 6 months after vaccination. We also show that only one dose of BNT162b2 is enough to achieve the maximum response in seropositive pre-vaccination donors.

## INTRODUCTION

SARS-CoV-2 is the virus responsible for the current COVID-19 pandemic. Several vaccines have been quickly and effectively developed to resolve this pandemic (Baden et al., 2021; Polack et al., 2020; Voysey et al., 2021). More than 390 million doses have been administered only in the United States by the end of September 2021 (OurWorldInData.org) and studies to investigate the comparative degree of protection provided by the different vaccines still have to be carried out. It is currently accepted that protection correlates with the humoral response (Plotkin, 2010; Plotkin and Gilbert, 2012). Moreover it is much easier to measure antibody than cellular responses and both often correlate to some extent (Grifoni et al., 2020). The Spike protein (S), abundant in SARS-CoV-2 viral surface, is highly immunogenic and antibodies against S are already detected one week after infection and lasting one year or more (Feng et al., 2021; Long et al., 2020). The individual humoral response to the S protein is highly variable (Garcia-Beltran et al., 2021) and this makes evaluation of the humoral response highly dependent on the reliability and sensitivity of the detection method. ELISA or CLIA are the most frequently used methods for specific antibody quantification. Both rely on detection of few epitopes present in recombinant protein fragments generated in conditions that do not fully reproduce the native status of infected cells (Galipeau et al., 2020). We have recently developed a highly sensitive method to detect specific IgG1, IgA and IgM against native SARS-CoV-2 S protein based on flow cytometry (Horndler et al., 2021a), from now on named SARS-CoV-2 S Jurkat Flow-Cytometry Immunoassay (JFCI). The S protein is expressed in the viral envelope, as well as in the surface of the cells used in the JFCI method, as a trimer. Due to its high sensitivity, JFCI allows to detect specific anti-S antibodies present in blood samples that were undetected by other methods like ELISA or CLIA, which miss the quaternary structure of the S protein (Horndler et al., 2021a; Valdivia et al., 2021).

In this study we have applied the JFCI method to carry out a comparative analysis of humoral SARS-CoV-2 S-specific immune response in volunteers that have been vaccinated with ChAdOx1 (Oxford-AstraZeneca), mRNA-1273 (Moderna) or BNT162b2 (Pfizer-BioNTech). We have found that although the 3 vaccines elicited a detectable humoral response in all blood donors after complete vaccination, there are quantitative differences both in serum IgG1 and IgA. In addition, we found that the magnitude of the antibody response declines with the age of the donor although it lasts up to at least 6 months post-vaccination. Finally, we have found that one dose of BNT162b2 is sufficient to achieve the maximum humoral response in seropositive pre-vaccination donors.

## RESULTS

We have developed a variant of the JFCI method to detect antibodies against the Spike protein of SARS-Cov-2 (Horndler et al., 2021a) where GFP, rather than truncated EGFR, is used as reporter protein. The human T-lymphoblastic Jurkat cell line was transduced with a lentivral vector that allows stable and coordinated co-expression of native S and GFP proteins from a monocistronic mRNA (Fig. 1A). For each individual test, Jurkat-S-GFP cells were stained with a 1:50 dilution of serum sample followed by anti-human IgG1 PE, anti-human IgM Pacific blue and anti-human IgA AF647. Dead cells were excluded with the viability dye 7AAD. Pools of positive and negative sera were used as controls to obtain two normalized values for each Ig isotype, both based on the fluorescence of Ig anti-S and GFP: score and ratio (Fig. 1B). The score is calculated by applying an algorithm based on the proportional distribution of both fluorescences and stablishes a cut-off to discriminate positivity (0.095 for IgG1, 0.081 for IgA and 0.11 for IgM). The main contributor to this value comes from the slope of the linear adjustment of the fluorescence intensities of anti-S and GFP, which is positive for a seropositive sample (blue line in contour plots in Fig. 1B). The Ig anti-S to GFP mean fluorescent intensity normalized ratio is used as a relative quantitative value as it correlates with the titer and affinity of specific antibodies to S (Horndler et al., 2021a). A Contour plot of the resulting staining with positive and negative control sera as well as the score and ratio values are shown in figure 1B as a representative result.

**Figure 1.**
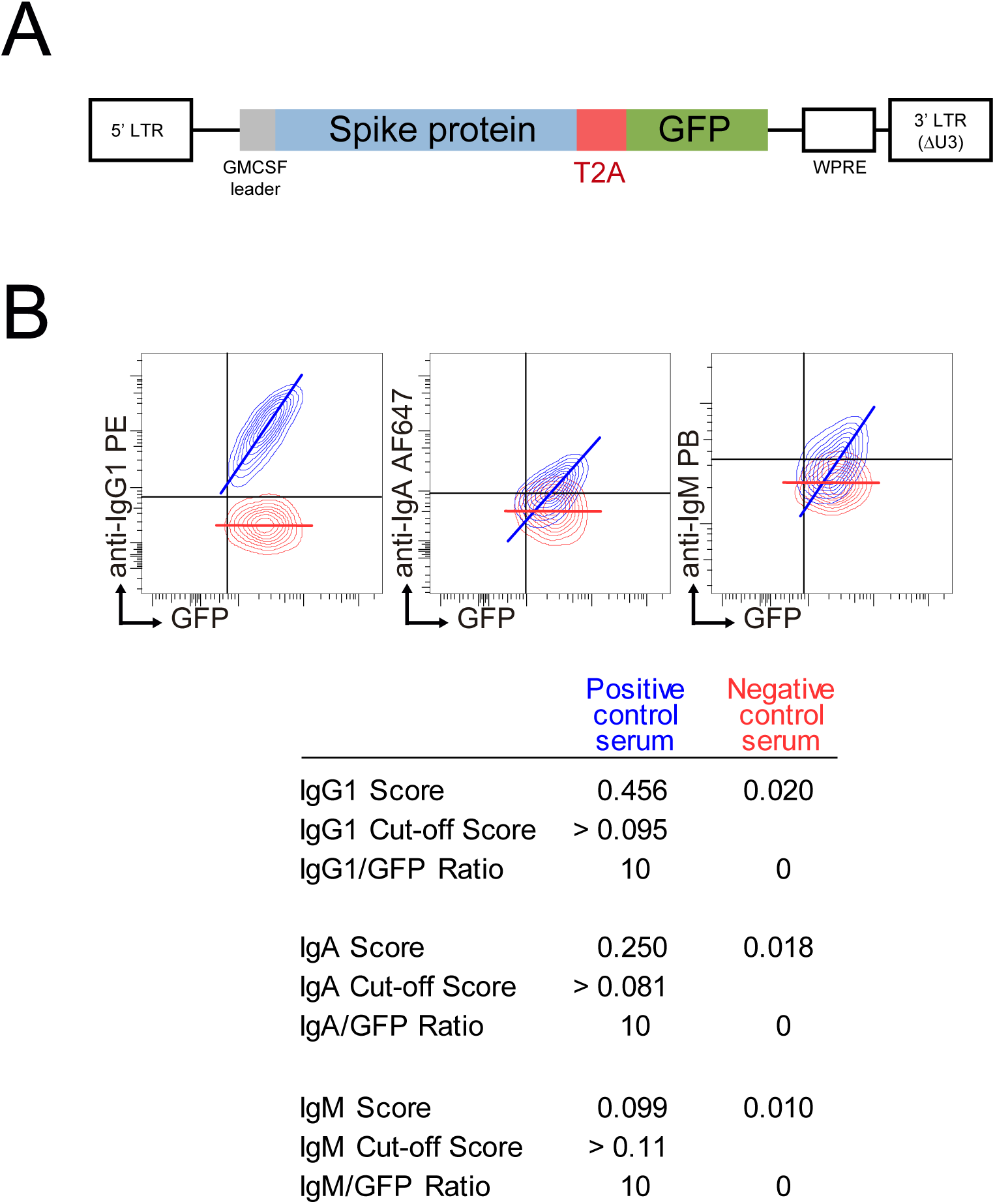
SARS-CoV-2 S Jurkat Flow-Cytometry Immunoassay (JFCI) method. **(A)** Lentiviral construct used to generate Jurkat S GFP cells. Full length mature SARS-CoV-2 Spike protein is followed by the T2A processing viral sequence and GFP. **(B)** Overlay contour plots of Jurkat S GFP cells stained with positive (blue) and negative (red) pool of sera. GFP and the three isotype signals are depicted. Score, cut-off value and normalized ratio for each sample and isotype are indicated in the table below.

Using this method, we have performed a comparative analysis of humoral SARS-CoV-2-specific immune response in ChAdOx1 (Oxford-AstraZeneca, ChAd), mRNA-1273 (Moderna, MO) and BNT162b2 (Pfizer-BioNTech, BNT) vaccinated individuals. A first cohort was composed of a total of 682 individual samples within which 59 ChAd, 36 MO and 165 BNT vaccinated donors at different time-points (Suppl. Table). Reported seropositive donors previous to vaccination were not included in this analysis. All donors became IgG1 seropositive after the vaccination was completed (post-dose 2, PD2) and seroconverted as soon as 3 weeks post-dose 1 (PD1) with the three vaccines (Fig. 2). Only a significant reduced proportion of seroconversion was detected after 2 weeks PD1 for BNT with 80% IgG1 seropositivity. In terms of potency, ChAd vaccine generated the lowest IgG1 titer, followed by BNT, and being MO the highest, since significant differences in IgG1 ratios were detected among the three groups at the comparable time-point group of 8 weeks PD2 (Fig. 3). Likewise, MO vaccine induced the highest IgA seroconversion with 96% seropositivity at 2 weeks PD1 compared to 67% and 74% for ChAd and BNT respectively (Fig. 2). It should be noted that not all ChAd and BNT donors became IgA seropositive at PD2. ChAd induced the lowest IgA titers, whereas MO and BNT reached similar values (Fig. 3). Regarding IgM, barely 11% of those vaccinated with BNT resulted positive at 8 weeks PD2 (Fig. 2), being this isotype the less effectively detected reporting the lowest titers (Fig 3) in all the groups.

**Figure 2.**
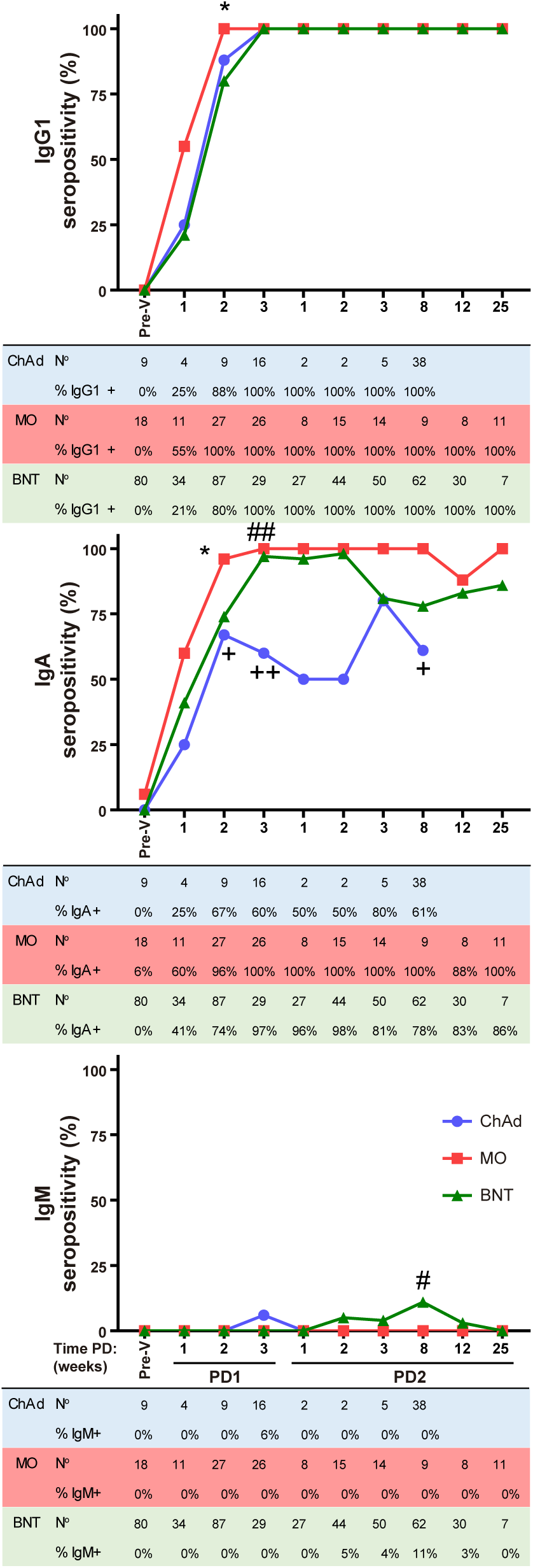
Ig anti-Spike seropositivity follow-up after vaccination. IgG1, IgA and IgM seropositivity in ChAdOx1 (ChAd), mRNA-1273 (MO) and BNT162b2 (BNT) vaccinated donors evaluated with the JFCI pre-vaccination (Pre-V) and at different time periods after the first (PD1) or the second (PD2) dose of the vaccines. Percentage of positive and total number of samples are depicted in tables below graphs. Samples were collected as 1 wk, 1-7 days; 2 wk, 8-16 days; 3 wk, 17-28 days; 8 wk, 29-60 days; 12 wk, 61-90 days; 25 wk, 91-174 days. Statistic comparisons were carried out for matched me PD between vaccines: +, ChAd vs MO; #, ChAd vs BNT; *, MO vs BNT.

**Figure 3.**
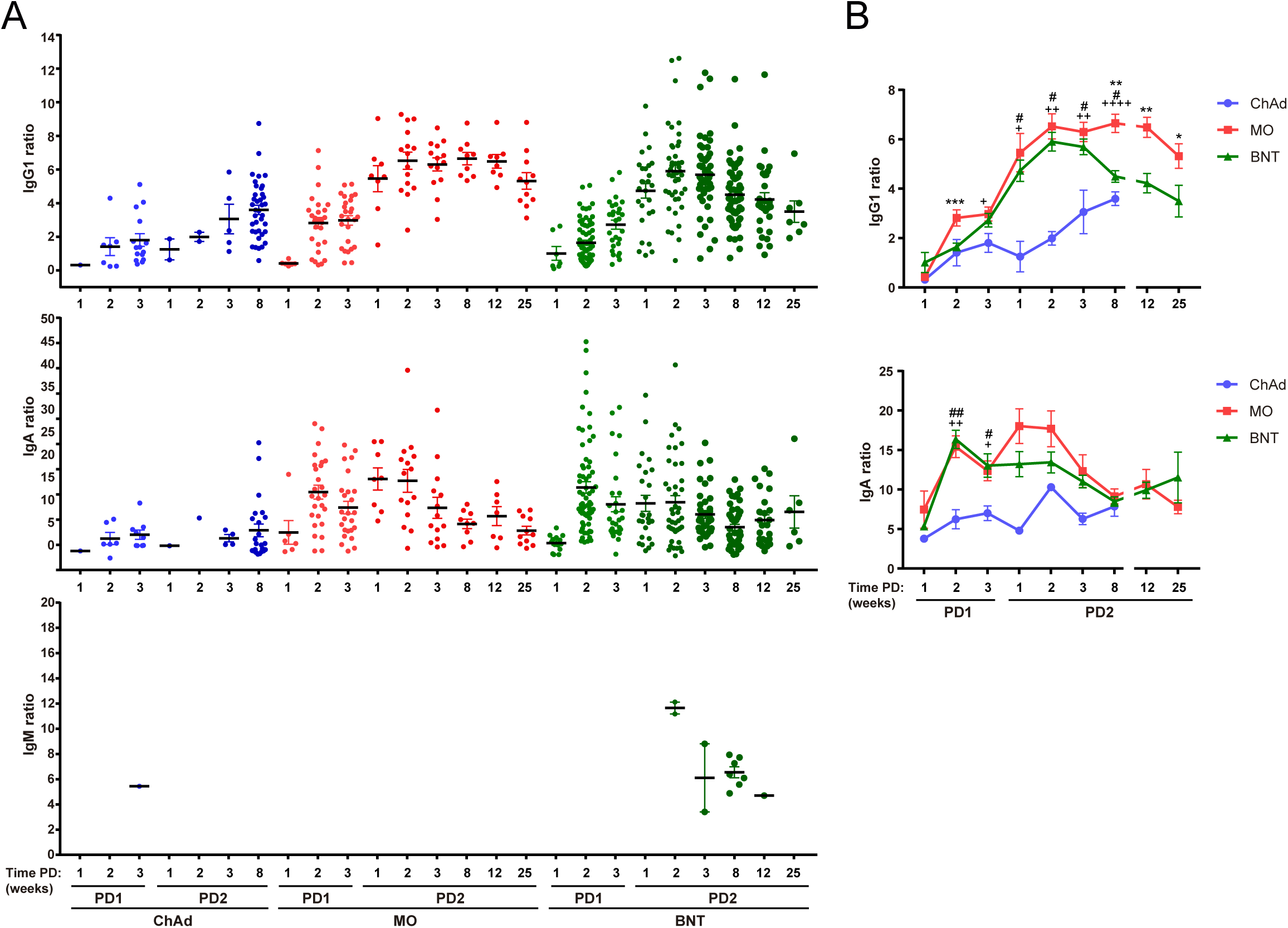
Ig anti-Spike titer follow-up after vaccination. **(A)** IgG1, IgA and IgM titters of seropositive samples described in figure 2, expressed as Ig to GFP ratio. **(B)** Overlay representation of data showed in A. Statistic comparisons were carried out for matched time PD between vaccines: +, ChAd vs MO; #, ChAd vs BNT; *, MO vs BNT.

In terms of kinetics, IgG1 titer increased at PD2 compared to PD1, reaching its maximum level at 3 weeks after the second dose for MO and BNT vaccines and declined in a slower fashion compared to IgA that peaked at 2 weeks PD1 for MO and BNT, declined and reached a second peak at 1 week PD2 for MO and bit later for BNT (Fig. 3). In spite the reduction observed, both isotypes remained quite stable at least up to the latest timepoint analyzed (25 weeks for MO and BNT). ChAd showed delayed kinetics reaching the maximum IgG1 and IgA titers at PD2.

There was a significant difference in the age of donors at 8-25 wk PD2 between MO and BNT groups (Suppl. Fig 1). The BNT donor group, derived mostly from nursing homes, were older on average than MO for the PD2 time-frame, and this difference in age could have an effect in the magnitude of the humoral response. For this reason, we next analyzed the effect of the age of the donor at the time of vaccination on the humoral response. We studied this effect within the larger cohort of samples, BNT, in which there was also a broad representation of donor ages. Sample-data were pooled as PD1 or PD2 and grouped by age in 20 year-intervals. Similar proportion of individuals became IgG1 or IgA seropositive both at PD1 or PD2 independently of the age (Table 1). However, IgG1 titers were significantly reduced in older seropositive donors both after the first and after the second dose (Fig. 4). By contrast, IgA titers increased with age. Similar results were obtained when data collection was restricted to 2-3 wk PD to avoid a possible bias due to differences in time PD (Suppl. Fig 2). Interestingly, IgM seropositivity was only found within the groups of older donors and after the second dose of vaccine (PD2) (Table 1 and Fig. 4). In those donors IgM was not produced in detriment of IgG1 (Suppl. Fig. 3A). In fact there is a direct correlation between IgM and IgG1 but not between IgM and IgA titers for all the IgM seropositive samples (Suppl. Fig. 3B). We also tested whether the donor’s gender had an influence in the humoral response and no difference was observed between male and female donors (Suppl. Fig. 4).

**Table 1.**
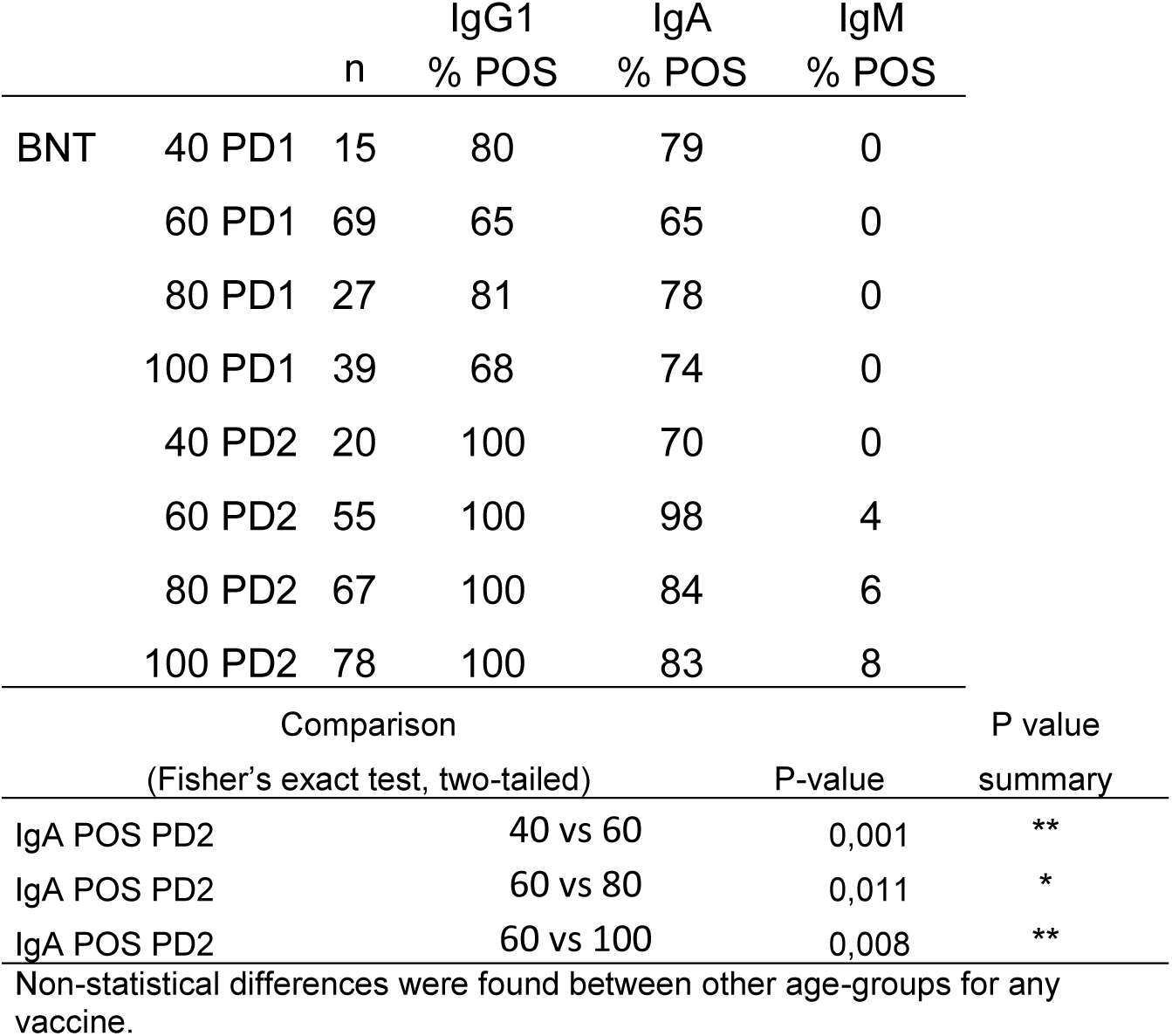
Total number and proportion of positive samples for IgG1, IgA and IgM after the administration of one or two doses grouped by the age of the donor.

**Figure 4.**
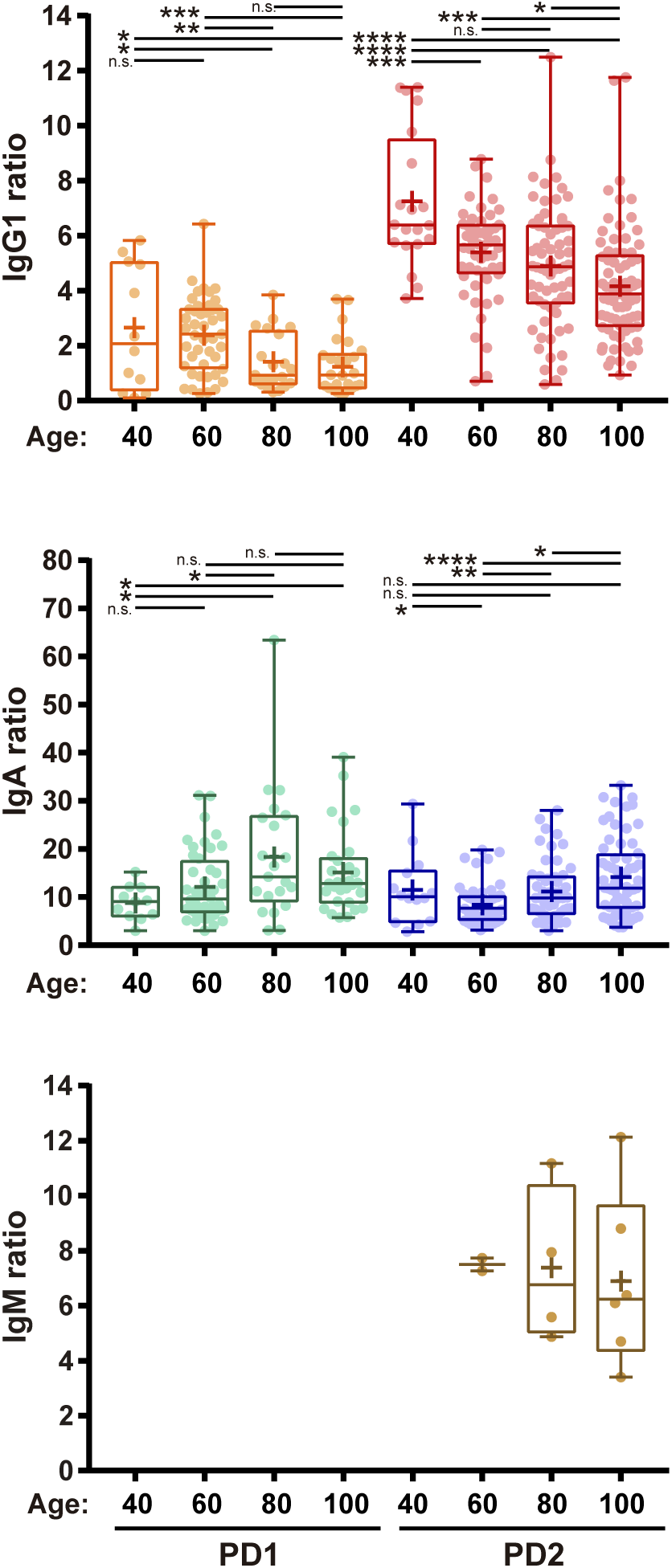
Effect of the age of the donor in the humoral respose. IgG1, IgA and IgM titers of seropositive samples collected after the first (PD1) or the second (PD2) dose of BNT vaccine. Samples were grouped by the age of the donor as 40, ≤ 40; 60, 41- 60; 80, 61-80; 100, 81-100 year-old. Median is showed inside boxes and Mean is represented with a + symbol.

A second cohort of 64 samples from 25 donors that had been previously infected with SARS-CoV-2 (and were IgG1 seropositive) prior to vaccination were evaluated after one or two doses of BNT compared to the first cohort of naive donors. We found that one dose of BNT administered to previously infected individuals was sufficient to induce the maximum level of IgG1 response (Fig. 5B). Furthermore, the humoral response did not increase after the second BNT dose in this group, unlike the group of naive individuals who had not been infected before vaccination. Nearly two-thirds of infected donors were already IgA seropositive prior to vaccination (Fig. 5A). The magnitude of the IgA response reached similar levels in both groups, although it was transiently sustained in previously infected donors decreasing to similar levels at 8 wk PD2 (Fig. 5B). Strikingly, IgM seropositive donors, although scarcely detected as occurred for the first cohort, were also detected in previously infected donor not only at PD2 but also at PD1 and pre-vaccination (Fig. 5A-B).

**Figure 5.**
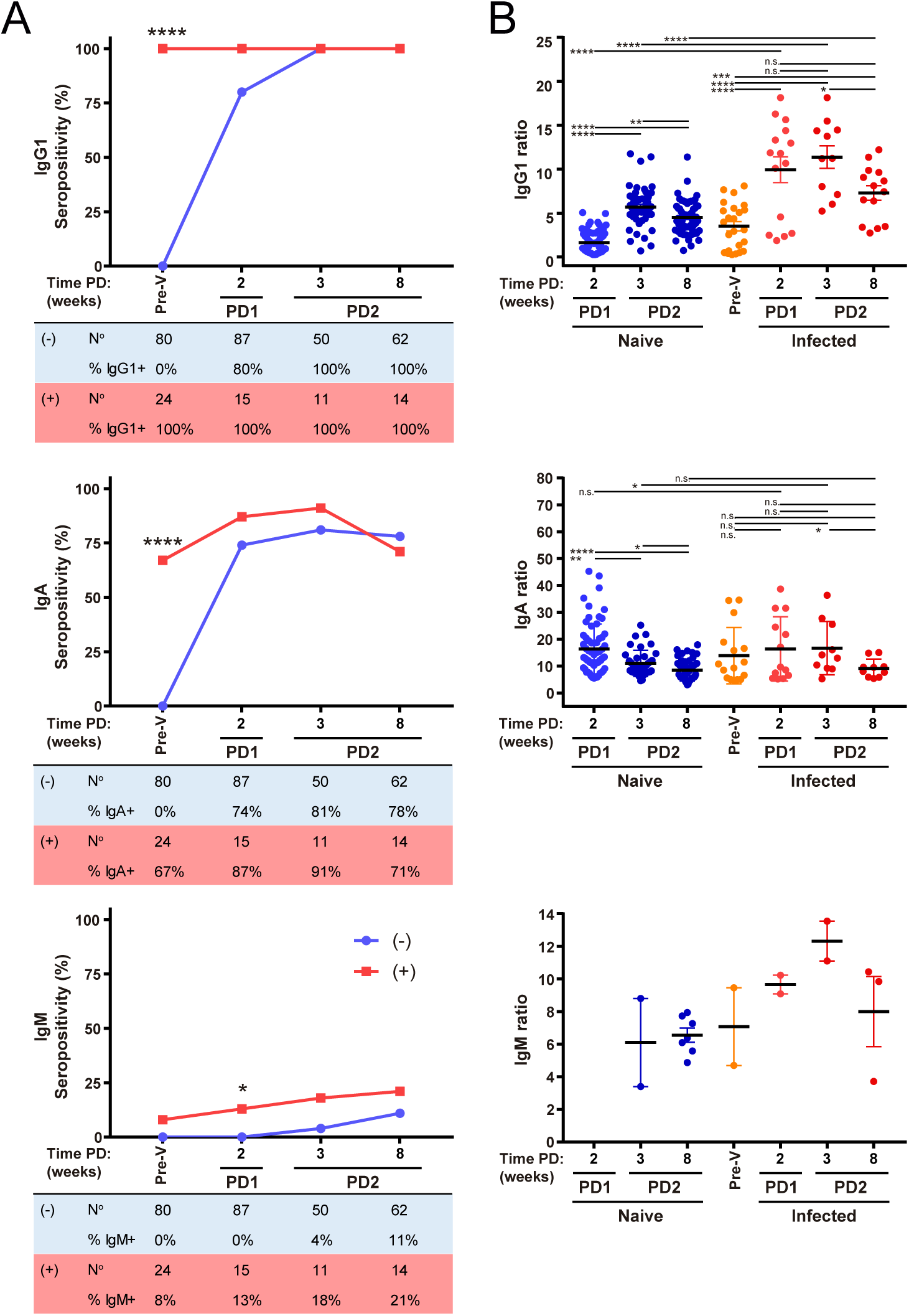
Humoral response in previously infected donors. IgG1, IgA and IgM seropositivity **(A)** and titers **(B)** of naïve (-) and previously infected and IgG1 seropositive (+) donors vaccinated with BNT. Total number and percentage of positive samples are shown in tables below seropositivity graphs. Sample collecon me as described in figure 2

## DISCUSSION

Here we have performed a longitudinal study to compare the humoral response to three different vaccines against SARS-CoV-2 in a cohort of Spanish individuals. We show that although the three vaccines eventually induced an efficient humoral response against the Spike protein of the virus, some differences in antibody seroconversion and titer exist. Seroconversion to IgG1 was fast with the three vaccines, reaching 100% seropositivity even only after one dose. All donors vaccinated with mRNA-1273 or BNT162b2 seroconverted to IgA whereas not all donors receiving the ChAdOx1 vaccine did. In the case of IgM only a minor seroconversion was detected in elderly donors vaccinated with BNT162b2. In terms of antibody titer, ChAdOx1 vaccine induced lower IgG1 and IgA titers whereas similar values were obtained for mRNA-1273 and BNT162b2. The differences in titers observed between the two mRNA vaccines could be age related (discussed below), as the mean age of donors for the BNT group is higher (Supplmentary Fig. 1). In summary our data indicate that mRNA-based vaccines, mRNA-1273 and BNT162b2, provoke a more potent humoral response and could be more immunogenic than the adeno-based ChAdOx1. Up to now only few studies have compared the humoral response of vaccinees adressed with identical test (Fabricius et al., 2021; Lechosa-Muñiz et al., 2021; Neumann et al., 2021): (i) Fabricius *et al*. found no difference in IgG and IgA titers PD1 between MO and BNT and increased IgA production at PD2 for MO. IgG and IgA titers were higher compared to ChAd but the latter received only one dose. Interestingly ChAd/MO and ChAd/BNT heterologus regimes for full vaccination induced higher IgG and IgA titers han ChAd/ChAd full vaccination. (ii) Lechosa-Muñiz *et al*. analyzed only one timepoint after full vaccination with mRNA vaccines or a single dose of ChAd and although they did not found a statistical difference between MO and BNT IgG titers, again they were higher compared to ChAd. (iii) Neumann *et al*. analyzed only IgG and found also a trend in higher titers for MO compared to BNT, PD1 or PD2. They also analyzed one dose for ChAd, giving the lowest titers. Our results are in accordance with these studies and expand the analysis to a complete vaccination regime.

The high sensitivity of the JFCI detection method allows not only the detection of low IgG1 titer but also the detection of IgA in serum. Most IgA antibodies are present in mucosal tissues. Total IgA is usually around 8 times (in average, it can be up to 40 times) less abundant in serum than IgG (total serum IgG range 7.5-22 mg/ml; total IgA range 0.5-3.4 mg/ml) and moreover usually only specific IgG and IgM are detected by ELISA/CLIA. It will be interesting to study if specific serum IgA detection could correlate with a proper IgA response in body cavities that would be essential to impede infection, prevent transmission or protect from disease severity. In fact IgA antibody responses have been detected in nasal fluids of patients infected with other coronavirus, and were associated with shortened periods of viral shedding (Callow, 1985). In this way serum IgA testing could serve as a correlate for protection.

We were not able to efficiently detect high levels of serum IgM. This could reflect a matter of antibody affinity. IgM is the first isotype rapidly secreted during the humoral response, mostly of low-affinity, leading to the production afterwards of high-affinity antibodies of different isotypes by the germinal center. The JFCI method is based on low protein expression on Jurkat cell surface, allowing the detection of high-affinity antibodies like IgG1 or IgA isotypes. On the contrary, ELISA is based on high concentration of recombinant protein fragments allowing the detection not only of high but also of low affinity antibodies. Higher epitope density likely allows for monogamous bivalent binding of low affinity antibodies. Other immunoassays to detect specific anti-Spike antibodies by flow cytometry have been developed and either do not show IgM detection (Hambach et al., 2021; Vesper et al., 2021) or they do efficiently detect IgM on beads where the antigen has been adsorbed (Cáceres-Martell et al., 2021; Dogan et al., 2021; Egia-Mendikute et al., 2021) or on the surface of HEK-293T cells where the antigen is overexpressed (Goh et al., 2021; Grzelak et al., 2020; Lapuente et al., 2021; Ng et al., 2020). In these two approaches epitope density is higher compared to the Jurkat-Spike cells of the JFCI, in line with our argument.

In terms of IgG1 and IgA kinetics our results show that humoral response to vaccination parallels the response to natural infection. It has been reported concomitant waves of IgA, IgM and IgG production in COVID-19 patients, being IgA and IgM cleared faster than IgG (Iyer et al., 2020; Sterlin et al., 2021). We have found an increase in IgA after each of the immunizations, reaching a peak around 2 weeks after priming or boosting immunizations and decreasing afterwards. On the contrary, IgG1 increased progressively reaching its maximum level around 2-4 weeks after the second boosting dose and decreasing slightly afterwards. Of note ChAd vaccine induced delayed kinetics, highlighting a decreased potency compared to mRNA vaccines. For both isotypes seropositivity is stabilized and maintained at least during the time period of this study, more than 6 months after the boosting immunization. Studies after this time frame are ongoing in order to know how long the response to vaccines persist. For IgM, as the JFCI does not allow detection of low affinity antibodies, we cannot compare responses to vaccines and natural infection, but it is interesting in any case as IgM produced at a long time after immunization could be of high affinity. Of note, humoral response in vaccinated individuals reached at least the same titer as natural infection.

Immunogenicity in the elderly has also been a matter of concern. The immune system efficacy is affected by the age, so a reduced humoral response could be expected in older vaccinated individuals. Seroconversion rate was not affected by age but we have found an inverse correlation between age and the magnitude of the IgG1 response. It would be also interesting to correlate the magnitude of the humoral response and protection since none of the individuals analyzed remained IgG1 seronegative after the complete vaccination. it is noteworthy that IgA titers, opposite to IgG1, increased with age and also that a few IgM seropositive cases were detected only after PD2 in older individuals as well as in the infected pre-vaccination group (in both cases the mean age of IgM seropositive individuals were 80 year-old). We did not find any relationship between IgM and IgA response (Suppl. Fig. 3B) and all IgM+ donors were also IgG1+ (Suppl. Fig. 3A). These observations reveal some kind of abnormality caused by age. A possible explanation could be a bias in isotype switching caused by alterations in the germinal center or an imbalance in long-lived plasma cells secreting IgA homing to mucosae. It could be also possible that memory against certain coronaviruses occurring only in the elderly could induce the generation of high affinity IgM. A decline in IgG antibody titers with age has also been observed after the second dose of mRNA vaccines (Goel et al., 2021). Although the cohort analyzed in that study was not enriched in subjects over 50, they also found a clear reduction in memory B cells with age. Other studies have reported a negative association between vaccine-induced antibody titers and age after a single dose of mRNA vaccines (Abu Jabal et al., 2021; Levi et al., 2021).

Finally, we show that previously seropositive individuals required only one dose of the vaccine to reach the maximum humoral response, reaching IgG1 greater titers than full-vaccinated donors. This result is in line with previous studies (Ebinger et al., 2021; Goel et al., 2021; Krammer et al., 2021; Stamatatos et al., 2021), and brings to light the importance of saving doses that would be unnecessarily administered to previously infected people. It would be more reasonable to reserve boosting to new versions of vaccines adapted to emerging variants of concern. But these decisions on vaccination should be taken with care because some people previously infected could mount a weak immunological response, especially those that do not develop COVID-19 symptoms, likely leaving these individuals with suboptimal protection. Diagnostic antibody testing would be required to take the more convenient decision, for which our JFCI would be of great usefulness. Moreover, serum IgG1 is the more informative isotype to test since it is the most persistent isotype present in blood.

Studies of the type herein described, analyzing longer times post-vaccination, and extended to other vaccines and vaccine-combinations, will be very useful to know the duration of the humoral response against SARS-CoV-2 in order to design the protocol for new boosting immunizations to sustain the response without wasting valuable doses. This will help to determine the better efficacy-cost ratio for a more suitable distribution of vaccine doses. In addition, prospective studies on vaccinated population testing not only humoral but also cellular responses will be required as a correlate for protection after infection with current and emerging viral variants. This will help to determine the degree of humoral response required for protection from infection and/or development of different degree of COVID-19 symptoms, a tool especially important to prove the efficacy of different vaccines and for the design of new vaccines.

## MATERIALS AND METHODS

### Cells

The human T-cell line Jurkat clone E6-1 was acquired from ATCC (TIB-152). Jurkat-S-GFP were stablished by transduction with the lentiviral vector based on the epHIV-7 plasmid where the human EGFR reporter was substituted by GFP and the full-length Spike S protein of Wuhan-Hu-1 was cloned (Horndler et al., 2021b). Cells were maintained in complete RPMI 1640 (GIBCO) supplemented with 10% fetal bovine serum (Sigma) and 100 U/mL penicillin-Streptomycin (GIBCO) in a humidified air-5% CO2 incubator at 37°C. Cells were routinely tested for the absence of mycoplasma.

### Blood samples and sera collection

Individual fingertip blood samples were taken in Microvette®200 Capillary Blood Collection tubes (Sarstedt). Then, sera collection was carried out after centrifugation at 10.000 rcf for 5 minutes, maintained at 4°C until use or -20°C for long time conservation. All participants provided written consent to participate in the study which was performed according to the EU guidelines and following the ethical principles of the Declaration of Helsinki. Serum sample collection was included in the study “ACE2 as a biomarker with utility for identification of high risk population for SARSCoV-2 infection and prognosis of evolution in COVID-19” approved by Autonomous University of Madrid Research Ethics Committee, no.2352. A description of samples within each cohort and full data is provided in supplementary Table.

### SARS-CoV-2 S Jurkat Flow-Cytometry Immunoassay (JFCI) procedure

Jurkat-S-GFP cells were adjusted to 1,2×10^5^ cells per well and plated in 96 well plates. Cells were centrifugated at 282 rcf for 2 minutes at 4°C and the supernatant was eliminated. The cell pellet was resuspended in 150 µl of working buffer that consist in phosphate-buffered saline (PBS), 2% bovine serum albumin (BSA, Sigma Aldrich) and 0,02% sodium azide (Sigma Aldrich) and washed by centrifugation. Then, cells were incubated with sera samples at a dilution of 1:50 with working buffer in a final volume of 100 µl for 20 minutes at 4°C. After incubation cells were washed to eliminate the unbound antibodies and possible interfering materials on the serum. Then cells were stained with 50 µl of the following cocktail of antibodies and viability reagent prepared in working buffer: 1:100 of mouse anti-human IgG1-PE (Ref.: 9054-09, Southern Biotech), 1:100 of mouse anti-human IgM-Pacific Blue (Ref.: PB-320-C100, Exbio), 1:50 of goat anti-human IgA-Alexa Fluor 647 (2052-31, Southern Biotech) and 1:50 of 7-AAD Viability Staining Solution (EXBOO26, Exbio). After incubation two additional washes were carried out. Cells were finally resuspended in the working buffer and analyzed on an Omnicyt™ Acoustic Focusing flow cytometer (Cytognos, S.L.) using the CytKick™ Autosampler (ThermoFisher Scientific). Data were processed with FlowJo software (BD). A pool of sera from 4 seronegative donors (unvaccinated and non infected) and other pool of sera from 5 seropositive and/or vaccinated donors were used as negative and positive controls respectively (Supplementary table). For each Ig isotype score cut-off values were determined with a collection of pre-COVID-19 sera and serial dilutions of seropositive samples and calculated as previously described described (Horndler et al., 2021a). Ig to GFP ratio was normalized to 10 for each isotype according to the positive control pool sera.

### Statistics

All data was analyzed using the GraphPad Prism 7 software. Outliers were removed from all data series with the Identify Outliers tool (Rout Q=0.1%). Error bars in figures represent SEM. Unpaired two-tailed Student’s t-test was applied to continuous data following a normal distribution and two-tailed Fisher’s exact test were used to assess differences between categorical variables. Differences were considered statistically significant at P≤0.05 (* p≤0.05; ** p≤0.01; *** p≤0.001; **** p<0.0001). Serum samples were received coded from the providers and the experimentalists were blinded to their nature until all data analysis was finalized.

## Data Availability

All data produced in the present work are contained in the manuscript

## DATA AVAILABILITY

This study includes no data deposited in external repositories. Jurkat-S cells are available for academic research upon request to B. Alarcón.

## Acknowledgments

We are indebted to all volunteers for generously participating in this study.

## Author contributions

SR-P, MQ and DA performed research and analyzed the data; AO and SA provided clinical samples, the original idea and edited the manuscript; BA analyzed data and supervised research, PD supervised and designed research, analyzed data and wrote the manuscript.

