## Supplementary Figures for "Longitudinal and comparative analysis of humoral response upon COVID-19 vaccination"

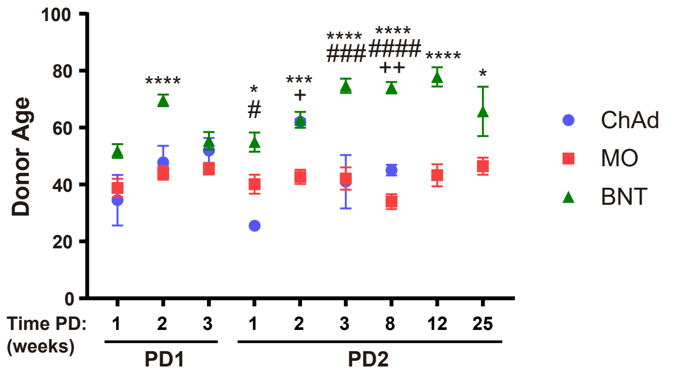


**Supplementary Figure 1. Donor age.**

Age of the donors analyzed in the first cohort of individuals vaccinated with ChAdOx1, mRNA-1273, and BNT162b2. Mean age for each timepoint group is represented. Statistic comparisons were carried out for matched time PD between vaccines: +, ChAd vs MO; #, ChAd vs BNT; *, MO vs BNT.

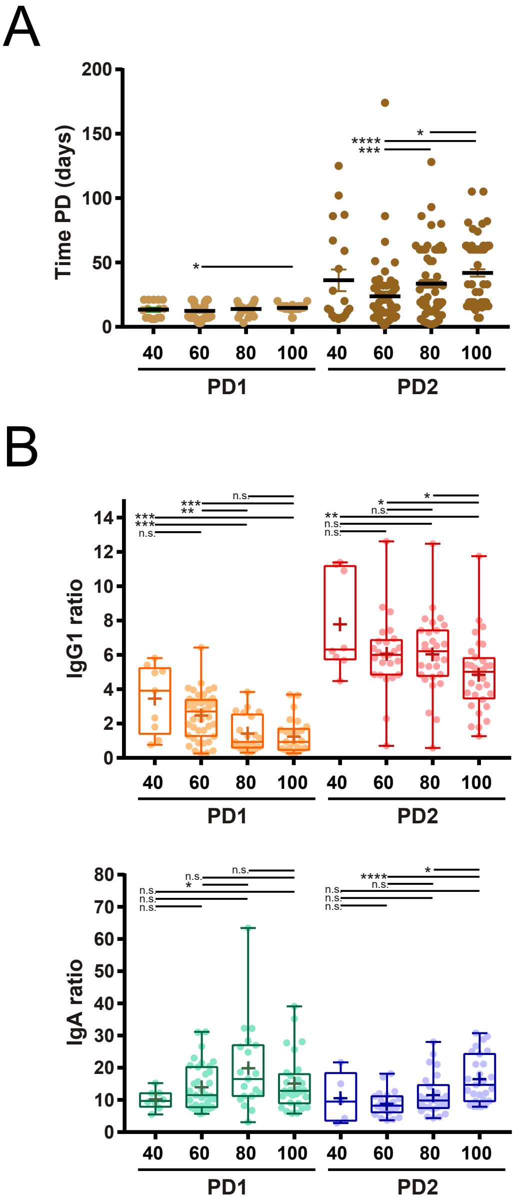


**Supplementary Figure 2. Time post-dose collection of samples related to the effect of the age of the donor in the humoral response.**

**(A)** Time post-dose collection of samples for the groups represented in figure 4.

**(B)** IgG1 and IgA titers of seropositive samples collected after the first (PD1) or the second (PD2) dose of BNT vaccine. Only 2 to 3 weeks PD samples were analyzed in the same way as for figure 4.

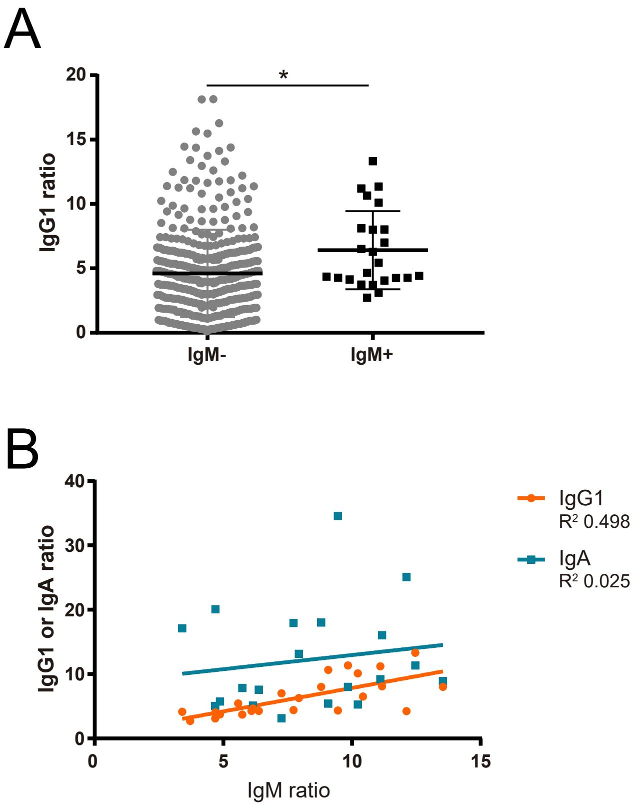


**Supplementary Figure 3. Relation between IgM and IgG1 or IgA titers.**

**(A)** IgG1 ratio for IgM seronegative (395) and IgM seropositive (24) samples from all BNT donors, naïve or infected pre-vaccination.

**(B)** Linear regression adjustment of IgM and IgG1 or IgA ratios for all IgM seropositive BNT donors (24), naïve or infected pre-vaccination.

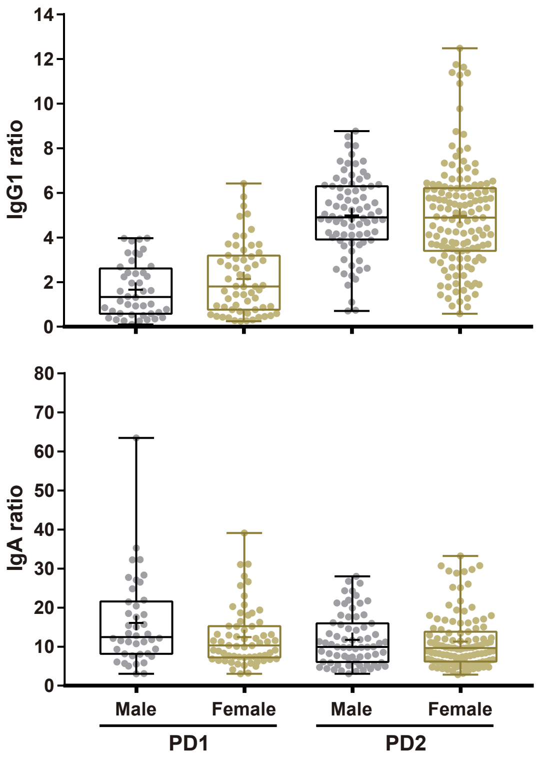


**Supplementary Figure 4. Effect the donor's gender in the humoral response.**
IgG1 and IgA titers of seropositive samples collected after the first (PD1) or the second (PD2) dose of BNT vaccine grouped by sex of the donor.
